# Exploring microbiota as a diagnostic tool for paediatric Ulcerative colitis and Crohn’s disease: a bioinformatics approach

**DOI:** 10.1101/2025.09.30.25336971

**Authors:** Xavier Tarragó, Alejandro Rodríguez, Javier Méndez, Javier Martín de Carpí, Gemma Pujol, Andreu Paytuví-Gallart, Antonio Monleón-Getino

## Abstract

Inflammatory bowel diseases (IBD), including Crohn’s disease (CD) and ulcerative colitis (UC), are chronic conditions that significantly impact paediatric patients. This study investigates differences in the expression of Kyoto Encyclopaedia of Genes and Genomes (KEGG) pathways and Gene Ontology (GO) terms between healthy individuals and patients diagnosed with CD and UC. The results reveal contrasting expression patterns between IBD patients and healthy individuals, suggesting differential regulation in pathways related to oxidative stress and inflammation, dysbiosis, alterations in protein synthesis and modification, energy pathways, homeostasis, and viral involvement. These findings provide new insights into the molecular mechanisms underlying IBD and highlight potential targets for future diagnostic and therapeutic strategies.

## 1. Introduction

Inflammatory bowel diseases (IBD) are a group of chronic disorders affecting the digestive system, characterised by symptoms such as diarrhoea, abdominal pain, rectal bleeding, faecal urgency, incontinence, and fatigue [1]. While the exact causes of IBD remain unclear, several contributing factors have been identified, including genetic predisposition, imbalances in the intestinal microbiota, environmental triggers, and alterations in the immune response [2]. IBD primarily encompass ulcerative colitis (UC) and Crohn’s disease (CD), which share some clinical features but differ in their location and effect on the gastrointestinal tract.

### 1.1 Evolution of inflammatory bowel diseases

Over the past five decades, the incidence and prevalence of IBD in Western countries have increased significantly [3]. The incidence per 100,000 population ranges from 8 to 14 cases for UC and from 6 to 15 cases for CD [3].

In Spain, Benallal et al. [4] analysed the incidence and characteristics of IBD in an area of Seville, comparing two time periods: 1995 - 2000 and 2001 - 2013. The results showed a significant increase in incidence, with new cases per 100,000 inhabitants per year rising from 4.2 in the first period (CD: 2.6; CU: 1.6) to 13.7 in the second period (CD: 7.2; CU: 6.5).

Up to 30% of IBD cases develop during childhood or adolescence [5], making early diagnosis essential to minimise disease impact on growth, nutrition, and pubertal development. Diagnostic criteria, such as the Porto criteria established by ESPGHAN, include complete colonoscopy, upper endoscopy with biopsies, and additional techniques such as ultrasound or magnetic resonance imaging [5]. Additionally, it is crucial to differentiate IBD from other inflammatory conditions and to address associated consequences, such as nutritional deficiencies and psychological effects, in a comprehensive treatment approach [5].

### 1.2 Differences between Ulcerative Colitis and Crohn’s Disease

UC manifests mainly in the colon and rectum, affecting the most superficial layer of the intestinal mucosa [6]. The inflammation in UC is continuous and often leads to the development of painful ulcers [7] Common symptoms include bloody diarrhoea, abdominal pain, and a general feeling of fatigue [6].

In contrast, CD can involve any part of the gastrointestinal tract, most frequently the terminal ileum and proximal colon [8]. Unlike UC, which is characterised by continuous inflammation, CD is marked by areas of inflammation interspersed with healthy tissue [8]. Symptoms of CD are similar to those of UC, and may include abdominal pain, bloody diarrhoea, weight loss, and persistent fatigue [6].

UC and CD are two of the main forms of IBD and both have a clinical course marked by alternating periods of exacerbation and remission [6]. During flare-ups, there is a significant increase in intestinal inflammation, resulting in the symptoms described above. In remission phases, inflammation decreases, and clinical symptoms significantly improve or disappear completely, although subclinical inflammation may persist in some patients [6]. The severity and duration of these cycles vary among individuals, often requiring a tailored therapeutic strategy to manage flare-ups and sustain long-term remission.

### 1.3 Impact of gut microbiota on inflammatory bowel diseases

The intestinal microbiota is a diverse community of microorganisms that live in symbiosis with the host and play an essential role in maintaining digestive health. Alteration of this balance, known as dysbiosis, has been linked with the development of gastrointestinal disorders such as IBD and colorectal cancer [9]. This dysregulation may compromise the integrity of the intestinal barrier, triggering persistent inflammatory processes and contributing to the development of pathologies [9].

According to a study by Nanjundappa et al [10], integrase produced by certain *Bacteroides* species can encode a mimotope of the pancreatic β-cell islet glucose-6-phosphatase catalytic subunit-related protein. A mimotope is a peptide that mimics the structure of an epitope native to the human body and is recognised by T cells. In this case, the mimotope activates CD8+ T cells implicated in diabetes. The T cells are recruited to the intestine, where they help suppress colitis-associated inflammation by targeting cells presenting the bacterial mimotope. This molecular mimicry mechanism, in which gut bacteria “mimic” host proteins, demonstrates how the microbiota can regulate immune activity. The study also suggests that these cytotoxic T cells may have therapeutic potential for the treatment of IBD [10].

The gut microbiota not only plays a fundamental role in digestive health but also interacts closely with the immune system to maintain homeostasis. When this balance is disturbed, the microbiota can trigger dysregulated immune responses, contributing to the development of IBD.

### 1.4. Objectives

This study aimed to identify molecular differences between healthy individuals and patients with CD or UC. Using data from faecal samples collected from paediatric patients at Sant Joan de Déu Hospital, we sought to analyse variations in gene expression, cellular interactions, and biochemical processes related to these pathologies. To achieve this, we employed Kyoto Encyclopaedia of Genes and Genomes (KEGG) pathways and Gene Ontology (GO) terms. We hypothesized that gene expression profiles would differ significantly between healthy individuals and those diagnosed with UC or CD, revealing distinct molecular signatures associated with each disease.

## 2. Methodology

### 2.1 Data Sources

The data used in this study were collected by Monleón-Getino et al. [11] from 32 paediatric patients at the Hospital Sant Joan de Déu. The participants were categorized into three groups: 13 patients with Crohn’s disease (CD), 11 with ulcerative colitis (UC), and 8 healthy controls (HC), with a follow-up period of 6 months (Table 1).

**Table 1.** Demographic characteristics of the experimental groups: ulcerative colitis (UC), Crohn’s disease (CD), and control) [11].

| Group | % Sex (male) | % Ethnicity (Caucasian, Arabian, Hispanic) | Age (Median, min, max) years |
| --- | --- | --- | --- |
| HC (controls) (*n* = 8) | 44.4% | 100%, 0%, 0% | 12 (10–16) |
| UC (*n* = 11) | 54.5% | 72.7%, 9.1%, 18.2% | 13 (9–15) |
| CD (*n* = 13) | 61.5% | 92.3%, 7.8%, 0% | 11 (8–15) |

Automated DNA extraction from oral and fecal samples was performed by Laboratorio Echevarne (Barcelona, Spain) following the manufacturers’ guidelines, as previously described by [11]. For oral samples, the QIAsymphony DSP Virus/Pathogen Kit (QIAGEN Iberia, S.L., Barcelona, Spain) was used in combination with the QIAsymphony SP system, while fecal samples were processed using the QIAamp PowerFecal Pro DNA Kit (QIAGEN Iberia, S.L., Barcelona, Spain) with the QIAcube Connect [11].

DNA-seq library preparation and sequencing were conducted by IGA Technology Services (Udine, Italy). The libraries for each sample were generated using the Nextera DNA Flex Library Prep Kit (Illumina, San Diego, CA, United States) following the manufacturer’s protocol, as detailed in [11]. Both input and final libraries were quantified using a Qubit 2.0 Fluorometer (Invitrogen, Carlsbad, CA, United States), and their quality was assessed with an Agilent 2100 Bioanalyzer High Sensitivity DNA assay (Agilent Technologies, Santa Clara, CA, United States). The libraries were then prepared for sequencing and processed on the NovaSeq 6000 platform in paired-end 150 mode (Illumina, San Diego, CA, United States) [11].

The primary bioinformatics analysis involved (1) base calling and demultiplexing, converting raw data format and de-multiplexing using Bcl2Fastq version 2.20 (Illumina, San Diego, CA, United States, RRID: SCR_015058); and (2) adapter sequence removal from raw fastq files using Cutadapt version 1.11, following the methodology outlined in [11].

### 2.2 Metagenomic analysis

The sequencing data initially underwent a quality check and trimming step with BBDuk with a minimum Phred quality score of 25 and minimum length of 35 base pairs. High-quality sequencing reads were aligned against the UniRef90 protein database using DIAMOND blastx [12], with a minimum read coverage threshold of 95% and a minimum identity of 80%. To resolve cases where reads aligned to multiple protein references, we employed a modified version of FAMLI [13] to assign each read to a unique protein. Gene Ontology (GO) terms and pathway annotations were then retrieved by mapping UniRef90 identifiers to the UniProt database via the UniProt API.

### 2.3 Clinical course of Crohn’s disease and ulcerative colitis treatment

Effective management of CD and UC requires monitoring of inflammatory markers and clinical responses to optimise therapeutic strategies. Data from the study by Monleón-Getino et al. [11] show that a large proportion of CD patients presented elevated inflammatory activity and abnormal inflammatory markers at diagnosis (Table 2). This was reflected in increased levels of faecal calprotectin (FC3) and elevated inflammatory markers such as erythrocyte sedimentation rate (ESR) and C-reactive protein (CRP) (Table 2). As treatment progressed, particularly with enteral nutrition, azathioprine, and biologic therapies such as anti-TNF agents, patients experienced significant clinical remission. This was reflected in the reduction of inflammatory markers at 3 and 6 months of follow-up and a decrease in FC3 levels (Table 2). At 3 months, only 33% of patients were classified in FC3, and at 6 months, this percentage had decreased further to 26% (Table 2). A similar pattern was observed in UC patients, who showed rapid improvement after treatment initiation. Clinical remission was achieved after 3 months of treatment and was maintained for 6 months in all but one patient (Table 2).

**Table 2.** Clinical data at diagnosis and follow-up (3 months and 6 months), including the paediatric Crohn’s disease activity index (PCDAI), paediatric ulcerative colitis activity index (PUCAI), faecal calprotectin (FC), erythrocyte sedimentation rate (ESR), and C-reactive protein (CRP). Patients were stratified into 3 subgroups based on FC levels: FC1 < 250 mg/kg (possible remission), FC2 250 mg/kg - 500 mg/kg (grey zone), and FC3 > 500 mg/kg (possible inflammatory activity) [11].

| Clinical group | Timeline | PCDAI/PUCAI mean (IQR) | FC (mg/kg) | FC stratified (%) |  |  | EIM (ESR and/or CRP) (%) |
| --- | --- | --- | --- | --- | --- | --- | --- |
|  |  |  |  | FC1 | FC2 | FC3 |  |
| CD | Diagnosis (n = 13) | 25 (10–47.5) | 2,166 (159–6,000) | 0% (n = 0) | 0% (n = 0) | 100% (n = 13) | 83% |
|  | 3 m (n = 12) | 0 (0–0) | 281 (50–2,880) | 50% (n = 6) | 17% (n = 2) | 33% (n = 4) | 0% |
|  | 6 m (n = 11) | 0 (0–0) | 93 (6–1,973) | 64% (n = 7) | 0% (n = 0) | 36% (n = 4) | 0% |
| UC | Diagnosis (n = 11) | 30 (10–80) | 3,381 (526–6,000) | 0% (n = 0) | 0% (n = 0) | 100% (n = 11) | 18% |
|  | 3 m (n = 10) | 0.5 (0–5) | 33 (7–1,042) | 90% (n = 9) | 0% (n = 0) | 10% (n = 1) | 0% |
|  | 6 m (n = 7) | 0.5 (0–5) | 9 (0–380) | 90% (n = 6) | 0% | 10% (n = 1) | 3.3% |

IQR, Interquartile range; PCDAI, Pediatric Crohn's Disease Activity Index; PUCAI, Pediatric Ulcerative Colitis Activity Index; FC, Fecal calprotectin; ESR, erythrocytic sedimentation rate; CRP, C-reactive protein; EIM, elevation of inflammatory markers.

### 2.4 Functional Analysis

To better understand how metabolic pathways and ontologies are affected by UC and CD and how these alterations may contribute to disease pathology, we used BIOFunctional, a program developed by our BIOST3 research group [14]. This tool, programmed in R, was specifically designed to analyse and visualise the complex relationships of KEGG pathways and GO terms in the different experimental groups at disease onset and during treatment (3 and 6 months).

BIOFunctional generates a directed acyclic network of gene ontologies and metabolic pathways across different disease stages (Fig 1). In this network, the nodes (circles) represent different disease stages, while the arrays (lines connecting nodes) indicate relationships between these elements. These relationships are classified according to their regulatory status: up-regulated (more expressed), down-regulated (less expressed), or neutral. In addition, each node and array is associated with a specific ontology or metabolic pathway, which allows the functional connections between the experimental groups to be identified (Fig 1).

**Fig 1.**
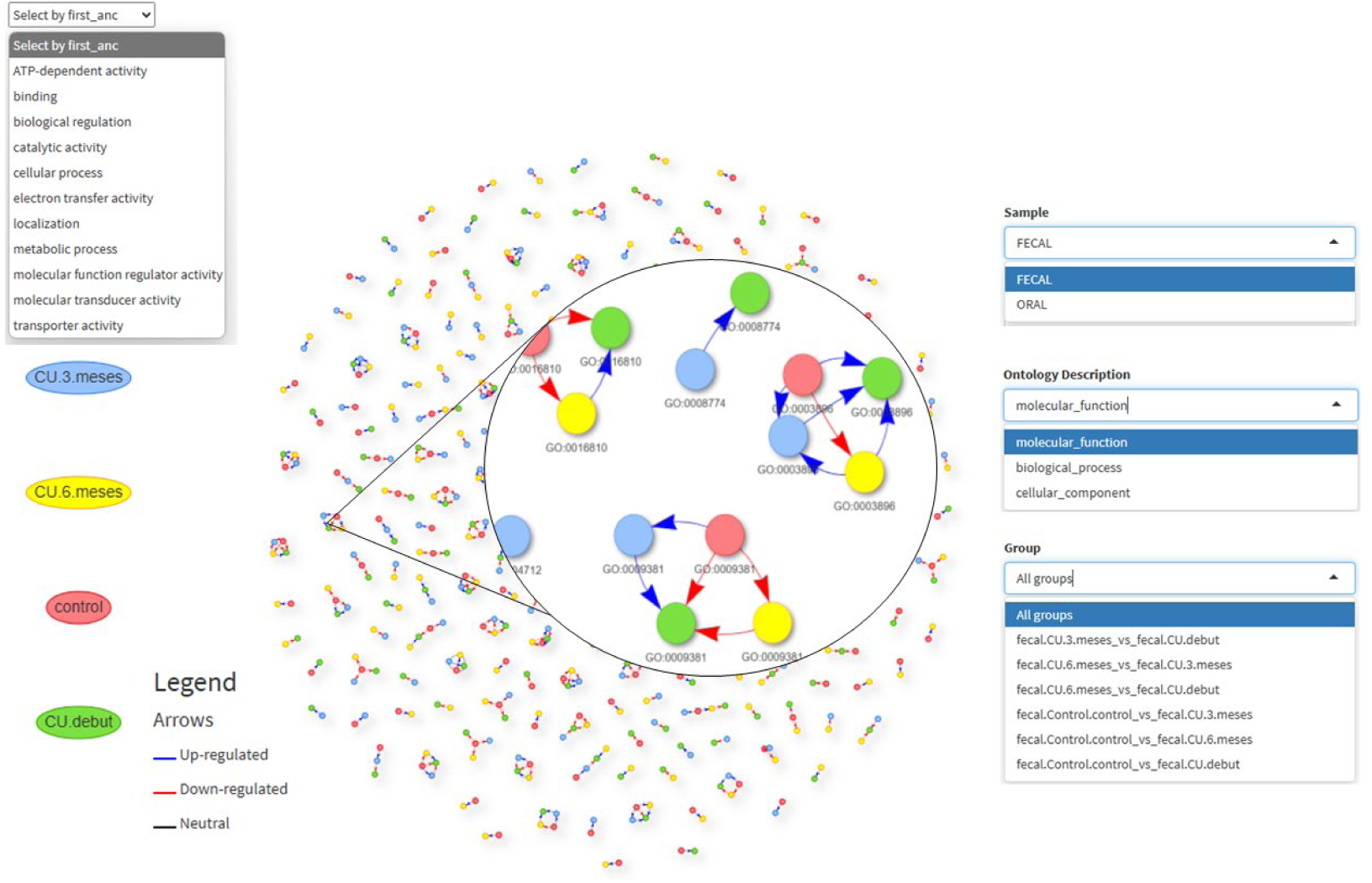
BIOFunctional biological interaction network overview. Visual representation of the BIOFunctional biological interaction network. Coloured nodes represent different experimental groups: red (control), green (onset), blue (3 months of treatment), and yellow (6 months of treatment). Unidirectional non-cyclic arrows indicate the connections and interactions between groups, with regulatory states represented by colour: black (neutral regulation), blue (up-regulation), and red (down-regulation). The top-right filter panel allows data selection according to categories such as “Molecular Function”, “Biological Process” and “Cellular Component”, which correspond to the three primary gene ontology domains. Additionally, it is possible to filter by sample type (faecal or oral). Within each domain, ontologies can be grouped according to the functions they share. It is also possible to filter by disease stages (e.g., control vs. onset).

The colour coding of nodes and arrays in the network is crucial for interpretation (Fig 1). Additionally, enrichment analysis (EA) was performed to determine whether a given pathway or ontology is overrepresented among the genes of interest beyond random expectation. The EA value is critical for assessing the biological relevance of a particular pathway or ontology in the context of IBD and facilitates the interpretation of functional changes associated with each disease state.

By applying this approach, we were able to identify differential patterns in metabolic pathways and ontologies associated with UC and CD. These findings provide new insights into the underlying mechanisms of these pathologies and their possible implications for disease progression and treatment.

## 3. Results and discussion

### 3.1 KEGG pathways in ulcerative colitis

Analysis of KEGG pathways in UC revealed distinct patterns of metabolic alterations at disease onset compared to the healthy control group. One notable change is a reduction in phenylpropanoid metabolism (Fig 2). Phenylpropanoids, obtained from a diet rich in plant-based foods, possess important antioxidant properties and contribute to maintaining the intestinal barrier [15]. According to Russell et al. [16], certain bacterial species in the colon can ferment aromatic amino acids to produce phenylpropanoid-derived metabolites. A decrease in phenylpropanoid metabolism may indicate dysbiosis, which can impair the ability of the gut to harness these protective compounds.

**Fig 2.**
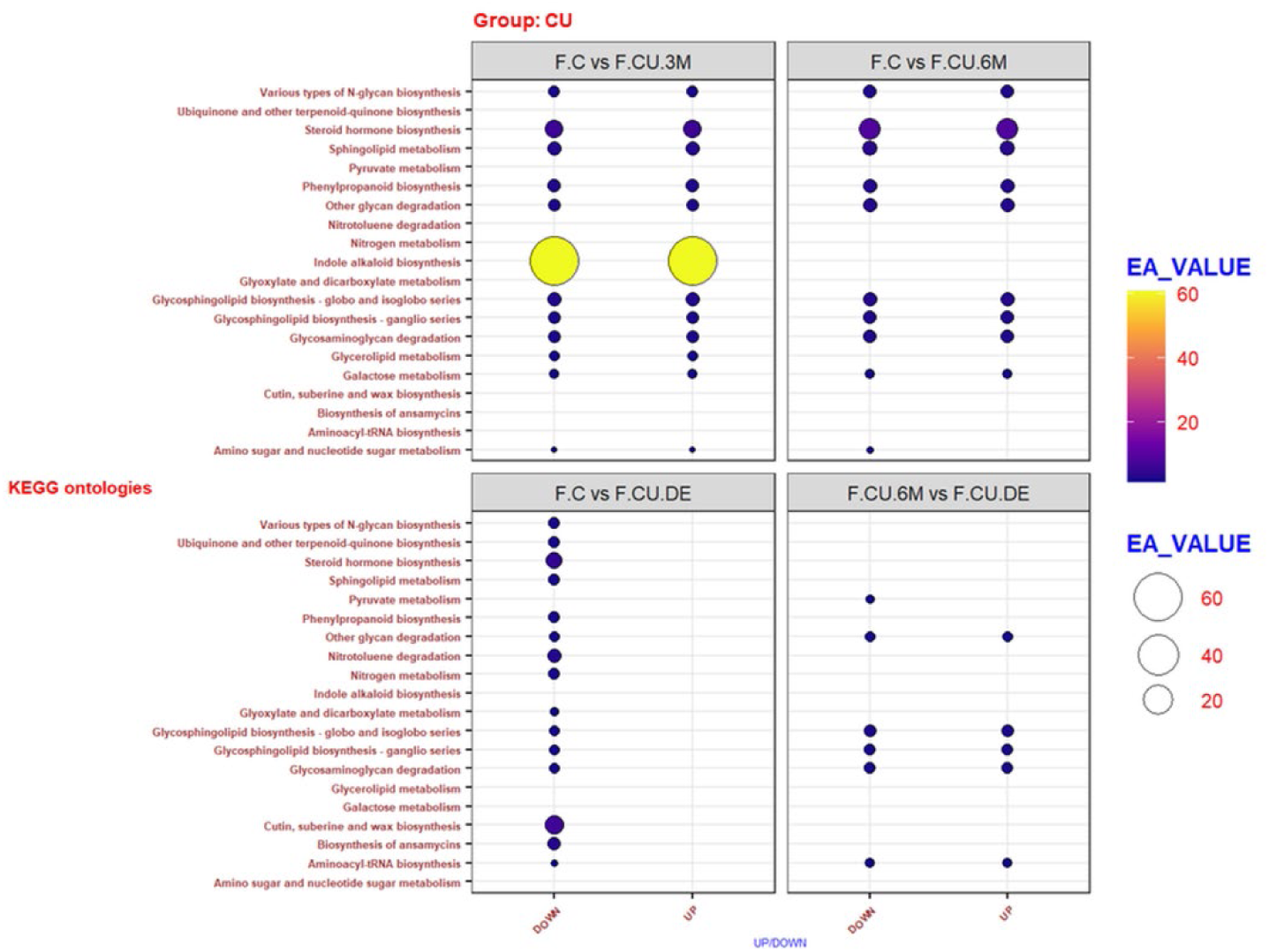
Enrichment Values across KEGG Pathways in faecal Samples from Ulcerative Colitis patients. Comparison of enrichment values (EA_VALUE) across KEGG metabolic pathways in faecal samples from ulcerative colitis patients, stratified by regulatory status (“Up” or “Down”). Each panel compares the control group with UC phases (onset, 3 months, and 6 months) and also compares the UC phases. Dot size and colour represent the level of enrichment, with larger dots and warmer colours indicating higher enrichment values.

Additionally, reductions were observed in the pathways of pyruvate, glyoxylate and dicarboxylate metabolism (Table 2). Pyruvate, a key intermediate in mitochondrial energy production, is converted to acetyl-CoA to fuel the Krebs cycle. Mitochondrial dysfunction, common in UC [17], can lead to inefficient energy production and exacerbate inflammation. Further research is needed to elucidate the underlying causes of the reduced activity in the glyoxylate and dicarboxylate cycles.

A decrease in glycan biosynthesis and degradation was also noted (Fig 2). Glycans play a key role in cell communication and immune response, and their alteration could impair inflammation control and tissue repair in UC [18]. They are involved in immunological processes such as antigen recognition, modulation of inflammation, and distinguishing between what is native or foreign to the body [18]. These functions are particularly relevant in inflammatory and autoimmune diseases such as UC, where immune regulation is often disrupted. Furthermore, glycans are essential for maintaining intestinal barrier integrity, as they contribute to the formation of mucins, which form a protective homeostatic barrier between the resident microbiota and the immune cells underneath the colon [19]. The observed decrease in glycan biosynthesis and degradation may have multiple causes and consequences, potentially leading to a vicious circle. A weakened intestinal barrier facilitates microbial contact with immune cells, triggering uncontrolled inflammation and contributing to UC pathogenesis and its characteristic intestinal dysbiosis.

Lipid metabolism pathways, including steroid hormone and sphingolipid biosynthesis, also show reduced activity at disease onset compared to healthy individuals (Fig 2). Glucocorticoids (GCs), which play a key role in regulating the inflammatory response, are naturally produced in the adrenal glands. The Hypothalamic-Pituitary-Adrenal (HPA) axis is activated when the body senses stress or inflammation. The hypothalamus secretes corticotropin-releasing hormone (CRH), stimulating the release of adrenocorticotropic hormone (ACTH) from the pituitary gland [20]. ACTH in turn prompts adrenal production of GCs, which exert anti-inflammatory effects [21]. GCs are a standard treatment for IBD, including CD and UC. They work by modulating the immune response and suppressing the production of pro-inflammatory cytokines such as interleukin (IL)-1, tumour necrosis factor, and IL-6, which are critical in the pathogenesis of the disease [21]. A reduction in GC synthesis may stem from negative feedback inhibition: chronically high levels of circulating GCs can suppress CRH and ACTH secretion, leading to lower GC production [20].

The observed decrease in glycosphingolipid levels at the early phase of UC may be related to alterations in lipid metabolism caused by intestinal inflammation. Gangliosides (glycosphingolipids consisting of a ceramide linked to fatty acids, sugar residues, and one or more sialic acids) play an important role in cell signalling and membrane maintenance. However, in IBD, these molecules undergo accelerated catabolism [22], which could impair their synthesis and contribute to the reduction of glycosphingolipids at this early disease stage.

Lower sphingolipid levels have also been associated with the fatigue commonly observed in IBD [23]. Additionally, the biosynthesis of ubiquinone, essential for the mitochondrial respiratory chain [24], is reduced in UC (Fig 2), likely due to intestinal dysbiosis and mitochondrial dysfunction [25]. Moreover, protein synthesis is compromised at the onset of UC, as evidenced by a decline in aminoacyl-tRNA biosynthesis [26] (Fig 2).

Taken together, these metabolic changes indicate a diminished capacity for antioxidant activity, energy production, immune regulation, and protein synthesis in the early stages of UC. In contrast, these pathways are more active during disease remission following treatment or in healthy individuals.

### 3.2 KEGG pathways in Crohn’s disease

KEGG pathway analysis in CD reveals distinct metabolic adaptations that vary according to disease stage and treatment. At disease onset, N-glycan biosynthesis is enriched (Fig 3), supporting a strong immune response, as N-glycans are essential for the functionality of antibodies and glycoproteins involved in inflammation [27]. However, this pathway stabilizes (becomes neutral) at 3 and 6 months of treatment (Fig 3). Alterations in N-glycan synthesis may contribute to chronic inflammation in inflammatory autoimmune intestinal diseases such as CD, as these molecules modify proteins involved in the response to inflammation and antigen recognition [27]. Additionally, N-glycans play a critical role in the modification of B and T cell receptors, which are responsible for antibody production and immune regulation. During the onset of autoimmune conditions, increased antibody production creates a higher demand for glycolysis of proteins involved in the immune response, such as cell-surface receptors and signalling molecules [27]. Moreover, many antibodies are glycoproteins that rely on N-glycans for stability and proper function. This could explain why the N-glycan biosynthetic pathway is more active at disease onset: the body is producing large quantities of antibodies to target tissues, and these need N-glycans to function effectively [27].

**Fig 3.**
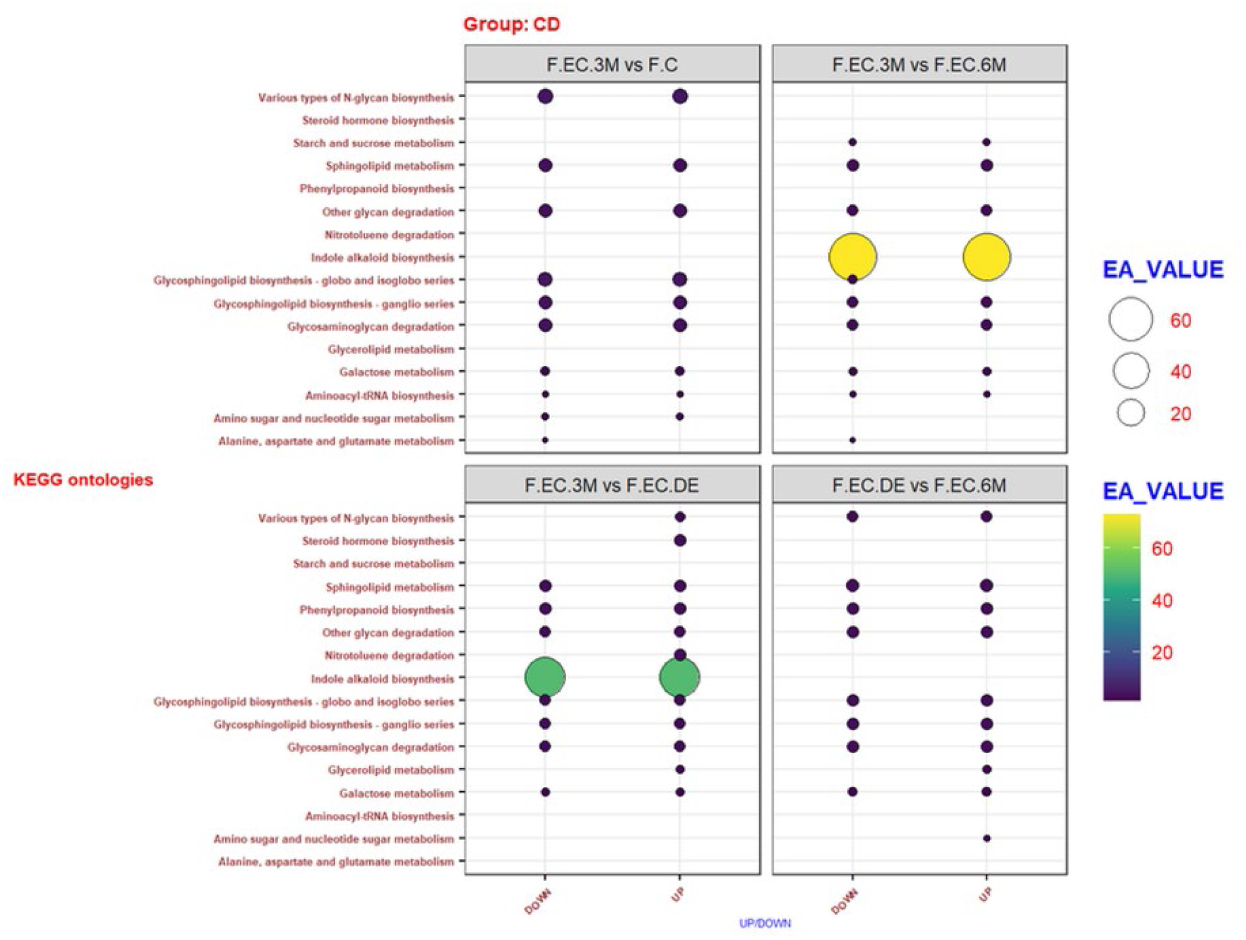
Enrichment Values across KEGG Pathways in faecal samples from Crohn’s Disease patients. Comparison of enrichment values (EA_VALUE) across KEGG metabolic pathways in faecal samples from Crohn’s disease (CD) patients, stratified by regulatory status (“Up” or “Down”). Each panel compares the control group with CD phases (onset, 3 months, and 6 months) or also compares the UC phases. Dot size and colour represent the level of enrichment, with larger dots and warmer colours indicating higher enrichment values.

An increase in glycerol metabolism and steroid biosynthesis are also apparent at disease onset (Fig 3). Glycerol serves as both an energy source and a precursor for lipids involved in the response to inflammation [28], while steroids exert anti-inflammatory effects [29]. After 3 months, glycerol metabolism was lower (Fig 3), suggesting that the treatment was moderating inflammation. However, it increased again at 6 months, possibly as part of an adaptive response, although the underlying mechanisms remain unclear (Fig 3). At 6 months of treatment, glycosphingolipid biosynthesis was lower than at 3 months (Fig 3), indicating a transition toward a more regulated immune system. Glycosphingolipids are key signalling molecules for immune cells such as T lymphocytes and dendritic cells [30], so their reduction may help mitigate chronic inflammation. In this phase of treatment, alanine, aspartate, and glutamate metabolism also decreased (Fig 3). During active disease episodes, these amino acids are often required for bioenergetic processes and protein synthesis associated with inflammation [31]. However, CD can lead to imbalances in amino acid metabolism, contributing to loss of muscle mass even during remission [31]. Although inflammation is reduced by treatment, persistent malabsorption of micronutrients in CD patients may impair amino acid utilization, limiting muscle protein synthesis.

Overall, CD disrupts metabolic pathways by impairing energy production, immune regulation, and anti-inflammatory processes, affecting both the gut microbiota and host metabolism. These changes include modifications in glycoprotein synthesis, lipid and amino acid metabolism, and cellular energy pathways, which adapt to the needs of the body in response to inflammation and treatment. These metabolic shifts can have significant consequences, including nutrient imbalances, chronic inflammation, and compromised immune function.

### 3.3 Differences in KEGG metabolic pathways between Ulcerative Colitis and Crohn’s disease

Although UC and CD are both forms of IBD with overlapping clinical features, they show distinct KEGG metabolic pathways. In UC, there is a decrease in glycan biosynthesis and degradation, as well as a reduction in steroid synthesis. These changes may be related to disruptions in the intestinal barrier and local immune regulatory mechanisms, which may contribute to the intestinal inflammation characteristic of UC. In contrast, the results for CD show an enrichment in both N-glycan biosynthesis and steroid synthesis. This increased metabolic activity may represent an adaptive response to chronic inflammation or indicate a distinct role for glycans and steroids in CD compared to UC.

The observed metabolic divergence between UC and CD is intriguing, given their clinical similarities. A more uniform metabolic response might be expected, particularly in key processes such as steroid synthesis or glycan regulation, which play critical roles in modulating inflammation and immune function. These metabolic differences may indicate fundamental variations in the pathophysiological mechanisms underlying each disease, although the precise biological implications remain unclear. Further studies are needed to understand why these metabolic processes differ so significantly and how these variations contribute to the distinct clinical manifestations of UC and CD.

**Table 3.** Summary of KEGG pathway differences between Ulcerative Colitis and Crohn’s Disease.

| **KEGG Pathway** | **Crohn's Disease (CD)** | **Ulcerative Colitis (UC)** |
| --- | --- | --- |
| N-glycan biosynthesis | Enriched at onset; stabilizes at later stages. | Decreased at onset. |
| Steroid biosynthesis | Increased at onset. | Decreased at onset. |
| Glycerol metabolism | Increased at onset, decreases at 3 months, then rises again at 6 months. | No significant change. |
| Glycosphingolipid biosynthesis | Decreased at 6 months. | Decreased at onset. |
| Alanine, aspartate, and glutamate metabolism | Decreased at 3 months, possibly linked to muscle mass loss. | No significant change. |
| Protein synthesis alteration | No significant changes. | Reduced aminoacyl-tRNA biosynthesis at onset, indicating impaired protein synthesis. |
| Energy pathways (pyruvate, ubiquinone) | No significant changes. | Decreased at onset, possibly linked to mitochondrial dysfunction. |
| Sphingolipid metabolism | No significant changes | Decreased at onset |

**Table 4.**
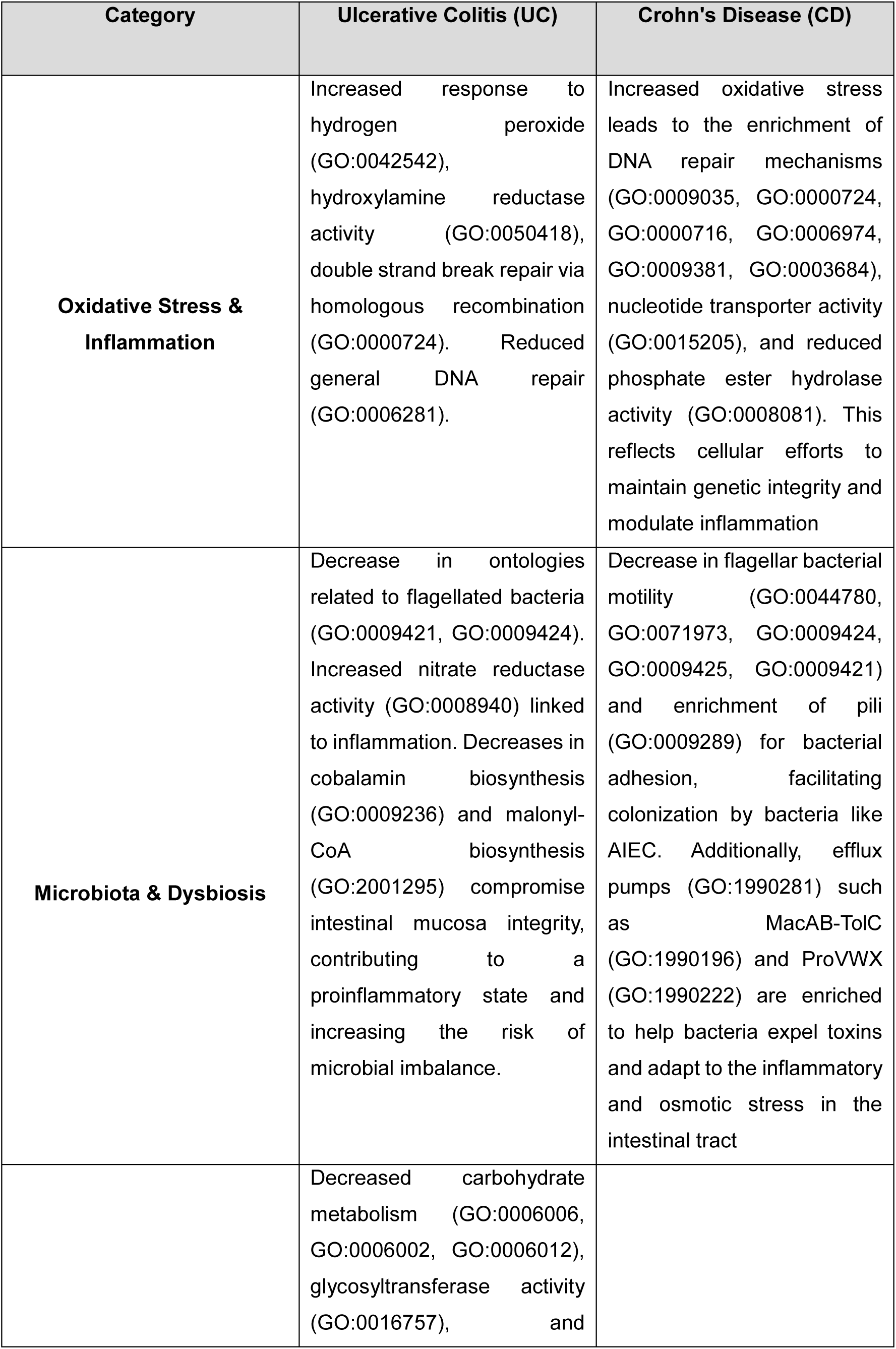
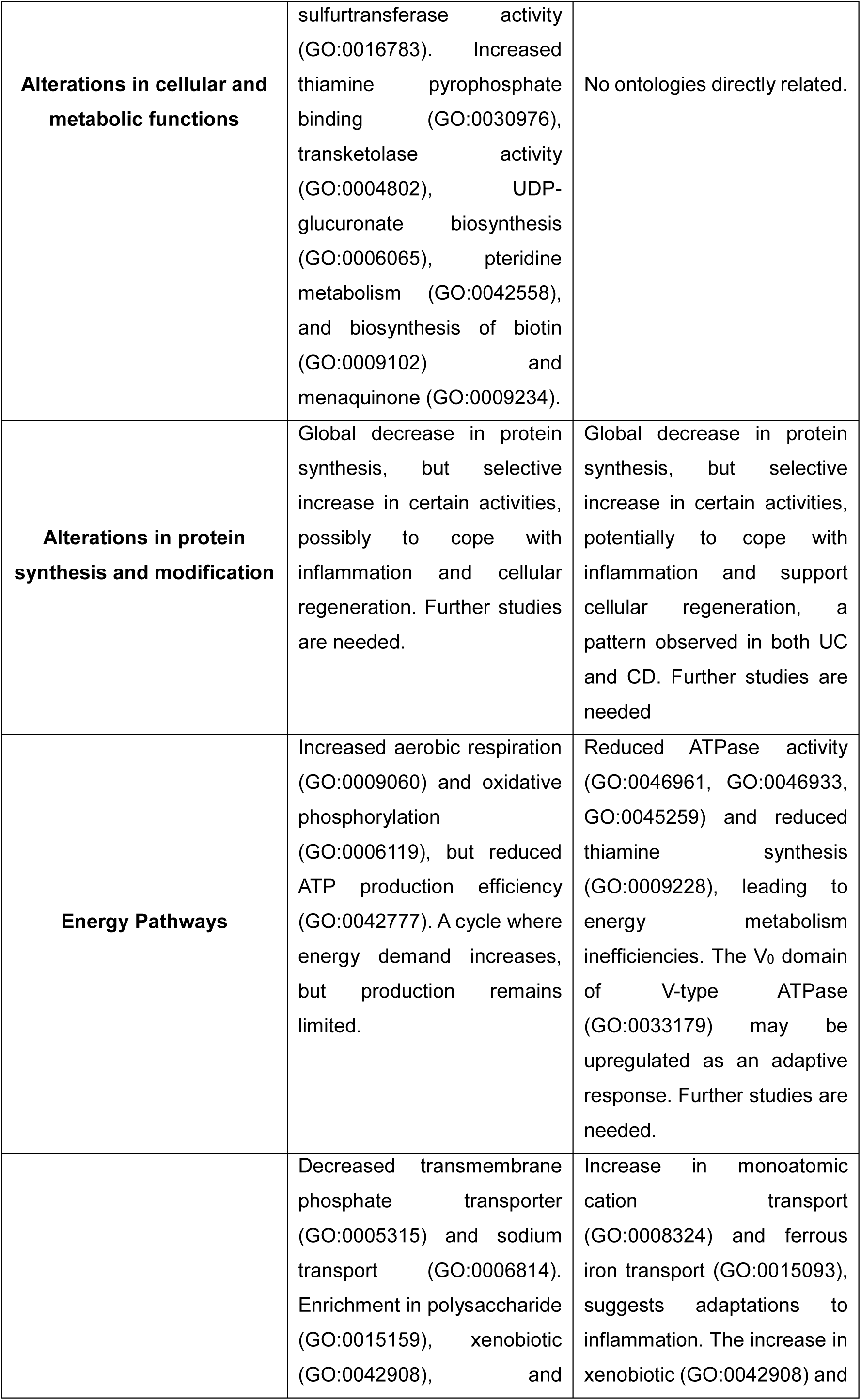
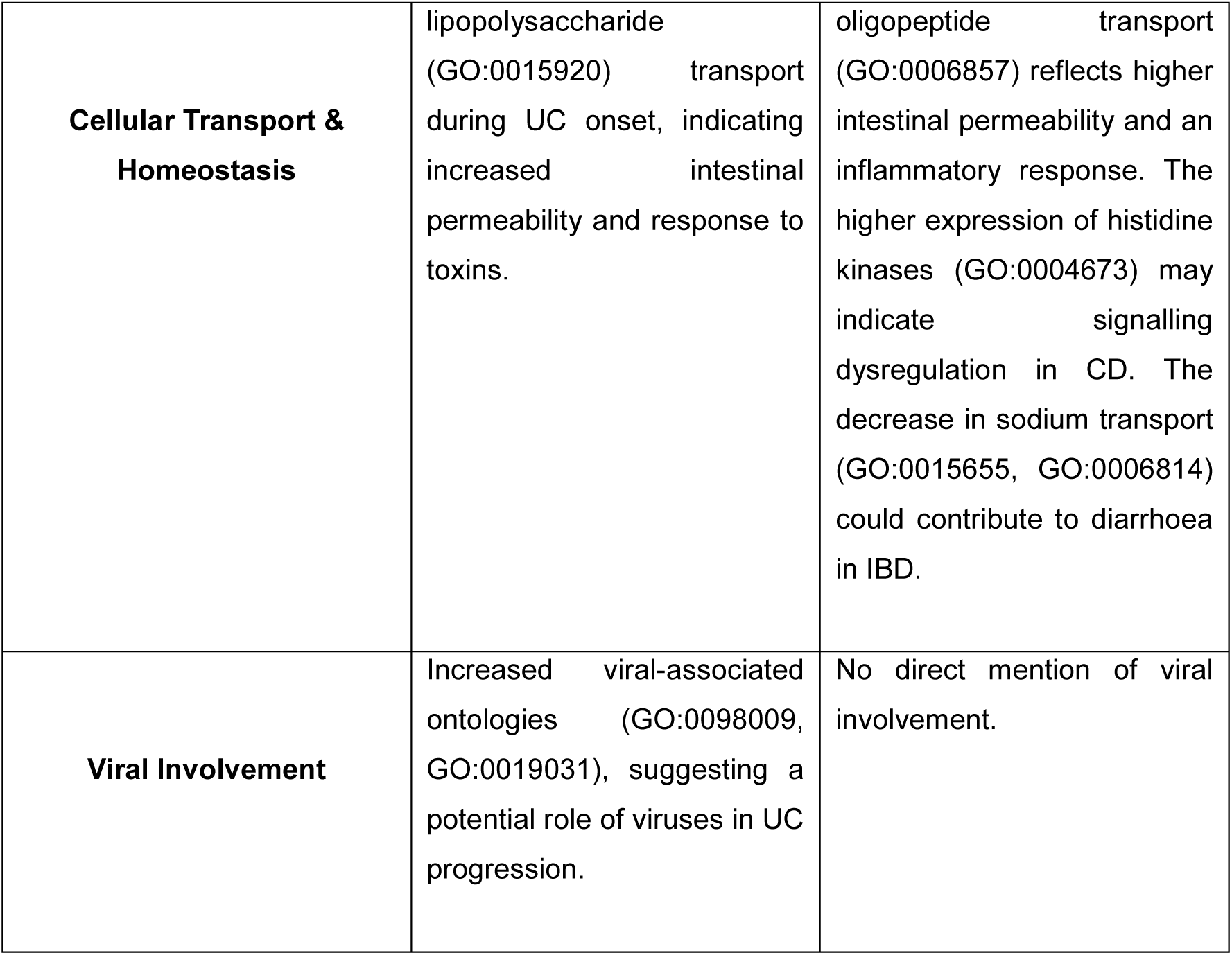
Summary of Gene Ontology key differences between Ulcerative Colitis and Crohn’s Disease.

### 3.4 Contextualizing gene ontology in Crohn’s Disease and Ulcerative Colitis

Gene ontology analysis in CD and UC provides valuable insights into the molecular and cellular changes associated with the progression of IBD. The findings reveal disruptions in various biological functions, metabolic processes, and signalling pathways associated with immune responses and gut microbiota, particularly during the early stages of both conditions. These alterations affect key processes such as metabolic regulation, signal transduction, cellular transport, energy pathways, and microbial dysbiosis. Understanding these changes can enhance our knowledge of the pathogenic mechanisms underlying CD and UC, as well as the adaptive responses of the intestinal system. Furthermore, these insights may contribute to the development of novel therapeutic strategies targeting the molecular pathways involved.

### 3.5 Gene ontology: Ulcerative colitis

To observe the differences in enrichment between the ontologies in the control group and the ulcerative colitis group, we generated bar charts displaying the up-regulated ontologies (Fig 4), covering the three main GO categories: Molecular Function, Biological Process, and Cellular Component.

**Fig 4.**
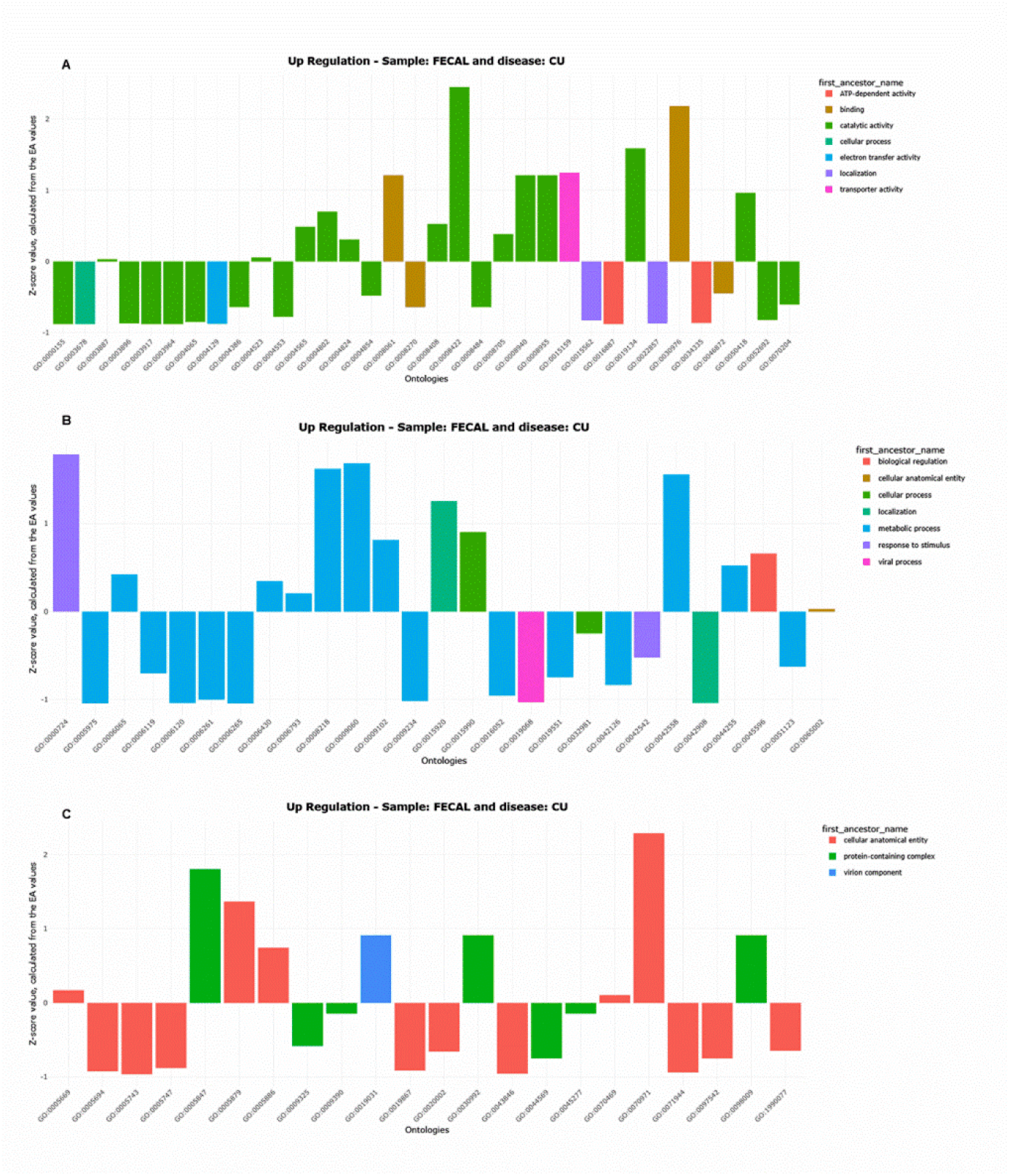
Bar Chart of Upregulated Ontologies in Ulcerative Colitis. Bar chart illustrating upregulated ontologies associated with ulcerative colitis (UC). Ontologies are plotted on the X-axis, and the Z-score (derived from EA values) is plotted on the Y-axis. The Z-score normalises enrichment values, indicating how far each ontology deviates from the mean within the same disease. This normalization helps identify the most relevant ontologies by highlighting those with significantly higher or lower enrichment. Fig A depicts molecular functions, Fig B biological processes, and Fig C cellular components, the three main gene ontology categories.

Similarly, the down-regulated ontologies (Fig 5) are also shown, following the same three GO categories.

**Fig 5.**
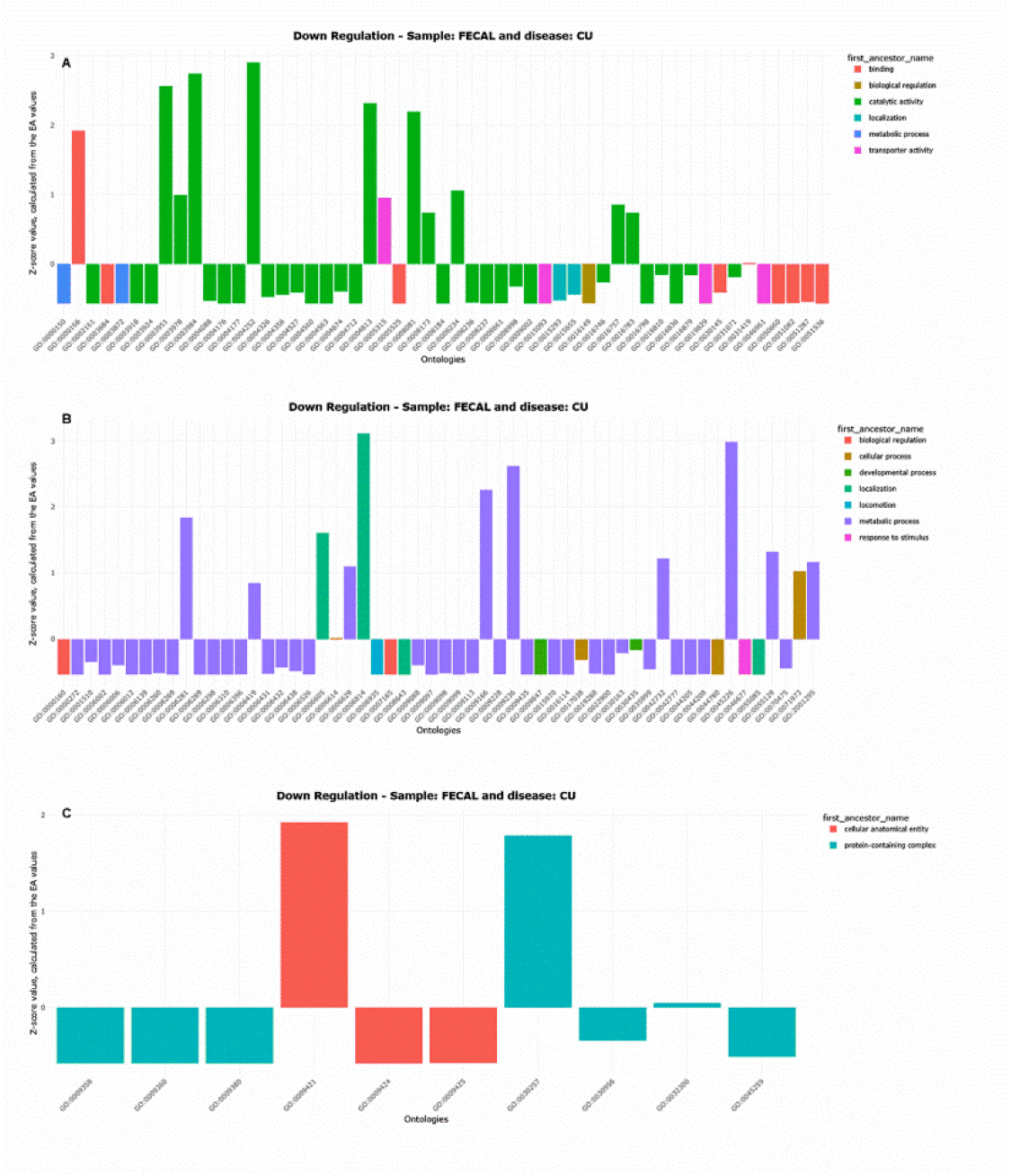
Bar Chart of Downregulated Ontologies in Ulcerative Colitis. Bar chart illustrating downregulated ontologies associated with ulcerative colitis (UC). Ontologies are plotted on the X-axis, and the Z-score (derived from EA values) is plotted on the Y-axis. The Z-score normalises enrichment values, indicating how far each ontology deviates from the mean within the same disease. This normalization helps identify the most relevant ontologies by highlighting those with significantly higher or lower enrichment. Fig A depicts molecular functions; Fig B, biological processes, and Fig C, cellular components, the three main gene ontology categories.

## 1. Response to oxidative stress and inflammation

Gene ontology analysis reveals a significant increase in pathways associated with the response to oxidative stress, particularly the response to hydrogen peroxide (GO:0042542) (Fig 4B). This suggests an activation of cellular mechanisms aimed at neutralizing reactive oxygen species [33]. Additionally, an increase in hydroxylamine reductase activity is observed (GO:0050418) (Fig 4A), which may be relevant in tissue damage and inflammation. This enzyme converts hydroxylamine into less toxic substances such as ammonia and water [34], helping to neutralise potentially harmful substances that accumulate during inflammation and disease progression [35].

This pattern is further supported by the enrichment of double-strand break repair by homologous recombination (GO:0000724) (Fig 4B), as well as DNA helicase activity (GO:0003678), 3’-5’ exonuclease activity (GO:0008408), and methionine synthase activity (GO:0008705), which are all involved in DNA repair. Methionine synthase plays a key role in folate metabolism and methionine synthesis, which are necessary for DNA methylation and the regeneration of damaged intestinal mucosa [36].

Conversely, a reduction in other DNA repair mechanisms (GO:0006281), nucleotide catabolism (GO:0009166) (Fig 5B), nucleotide binding (GO:0000166), and DNA strand interchange activity (GO: 0000150) (Fig 5A) suggests that during UC onset, specific repair processes are prioritized to address severe damage, such as double-strand breaks [37], rather than general repair pathways. This may be a consequence of intense oxidative stress, which induces more extensive damage, thereby triggering the homologous recombination pathway to preserve genetic integrity [38].

## 2. Dysbiosis and alterations in the microbiota

Dysbiosis is evident at the onset of UC, as indicated by the reduction in gene ontologies related to flagellated bacteria (GO:0009421, GO:0009424, GO:0009425, GO:0044780, GO:0071973) (Figs 5B and 5C). This decline suggests a decrease in bacteria with active flagellar motility, possibly due to inflammation creating an inhospitable environment. As reported by Hsu et al. [39], under inflamed conditions, gut cells secrete proteins such as hLYPD8, which bind to bacterial flagella, limiting their mobility and preventing invasion of the epithelial barrier.

The observed increase in nitrate reductase activity (GO:0008940) (Fig 4A) may be related to elevated nitric oxide (NO) production [40]. NO is a key mediator in inflammatory and autoimmune responses, with nitrate reductase catalysing the conversion of nitrates to nitrites, which can then be used for NO synthesis. NO plays a critical role in tissue destruction during inflammation, a process especially relevant to IBD pathogenesis [40]. Furthermore, Pool et al. [40] reported that serum nitrate levels are higher during active UC compared to the quiescent state, indicating that NO production increases during disease flares. This rise in nitrate levels in patients with active UC suggests a direct pathophysiological involvement of nitrate reductase in intestinal inflammation [40]. Additionally, the increased activity of peptidoglycan glycosyl transferase (GO:0008955) (Fig 4A), a key enzyme in bacterial cell wall synthesis, reflects an adaptive shift in the intestinal microbiota, possibly in response to changes in the intestinal environment [41].

Ontologies associated with lipid metabolism (GO:0006629) (Fig 5B), cobalamin biosynthesis (GO:0009236) (Fig 5B), and malonyl-CoA biosynthesis (GO:2001295) (Fig 5B) show lower enrichment at disease onset compared to the control group. The gut microbiota regulates several key metabolic processes for the host, including energy homeostasis, glucose metabolism, and lipid metabolism [42]. Microbial imbalance, a hallmark of UC, can disrupt these metabolic functions, leading to reduced activity in these pathways [42]. This disruption is evident in the decreased biosynthesis of malonyl-CoA, a key enzyme in lipid synthesis. Its deficiency limits the availability of essential lipid precursors required for cellular repair, thereby exacerbating inflammation and increasing the vulnerability of plasma membranes [43].

Likewise, reduced cobalamin (vitamin B12) production during the onset of UC may impair the ability of the intestinal mucosa to maintain its structure and function in the face of inflammation. Cobalamin, primarily produced by gut microorganisms, is vital for intestinal mucosal health [44]. A decline in its production, as observed in UC, can compromise mucosal integrity, exacerbate a proinflammatory state, and increase the risk of microbial dysbiosis or imbalance [44].

## 3. Alterations in cellular and metabolic functions

In early stages of UC, significant changes in carbohydrate metabolism are observed, as indicated by the reduced activity of pathways associated with glucose (GO:0006006), fructose (GO:0006002), and galactose (GO:0006012) metabolism (Fig 5B). This decline is likely a consequence of the inflammatory stress characteristic of UC, which impairs the ability of intestinal cells to efficiently use these nutrients [45].

In contrast, an increase was noted in the binding of thiamine pyrophosphate (GO:0030976) (Fig 4A), an essential cofactor for several enzymes involved in energy metabolism, especially glycolysis, the pentose phosphate pathway, and the Krebs cycle [46]. This adaptation may represent an attempt by cells to maintain optimal energy function by relying on pathways that are less susceptible to inflammatory or oxidative damage or that use different substrates.

Regarding the intestinal barrier, a reduction in enzymatic activities crucial for its integrity was observed, such as glycosyltransferase (GO:0016757) and sulfurtransferase activity (GO:0016783) (Fig 5A). This decrease is possibly due to the influence of proinflammatory cytokines, which modulate the expression of these enzymes [47]. The resulting impairment of glycosylation disrupts the function of glycoconjugates, which are essential for cell-cell interactions, intestinal barrier maintenance, and immune regulation [47]. Thus, dysfunction in glycosylation may exacerbate inflammation and disease progression by disrupting normal cellular functions.

Plasma membrane enrichment (GO:0005886) (Fig 5C) in UC is associated with structural and functional changes in the epithelial barrier. Chronic inflammation and dysregulation of key proteins compromise the integrity of the epithelial barrier. UC is characterized by intense mucosal inflammation in the colon, often leading to the loss of tight junction proteins such as zonula occludens (ZO) proteins, including ZO-1, ZO-2 and ZO-3, which are responsible for maintaining epithelial cell cohesion and membrane impermeability [48]. The disruption of the epithelial barrier increases membrane permeability, rendering it more susceptible to pathogenic factors and promoting chronic inflammation [48].

Additionally, transketolase (TKT) activity (GO:0004802), a key enzyme in the pentose phosphate pathway (PPP), is enhanced (Fig 4A). Increased TKT activity may have beneficial effects on certain aspects of the inflammatory response and tissue repair. According to Tian et al. [49], TKT serves as a guardian of intestinal integrity and barrier function, making it a potential therapeutic target for intestinal disorders. Inhibition of TKT leads to significant intestinal alterations, including extensive mucosal erosion, aberrant tight junctions, impaired barrier function, and increased inflammatory cell infiltration. Mechanistically, TKT deficiency results in the accumulation of PPP metabolites and depletion of glycolytic intermediates, thereby reducing ATP production, and triggering excessive apoptosis, ultimately comprising intestinal barrier function.

The upregulation of UDP-glucuronate biosynthesis (GO:0006065) and the metabolism of pteridine-containing compounds (GO:0042558) (Fig 4B) during the onset of UC reflects the inflammatory state characteristic of the disease. In particular, UDP-glucuronate biosynthesis is involved in detoxification and the response to exogenous compounds, essential processes in a proinflammatory environment [50]. Similarly, the metabolism of pteridine-containing compounds contributes to the synthesis of essential cofactors for enzymatic function, which is vital for maintaining cellular function under chronic inflammatory conditions [51].

Furthermore, an enrichment in the biosynthesis of vitamins such as biotin (GO:0009102) and menaquinone (GO:0009234) is observed (Fig 4B). These cofactors may play an important role in the adaptive response to inflammatory stress by modulating the immune response and preserving cellular function [52].

## 4. Energy pathways

The increased activity of energy pathways, such as aerobic respiration (GO:0009060) and oxidative phosphorylation (GO:0006119) (Fig 4B), reflects a cellular effort to meet the high energy demands associated with the immune response and tissue repair. Enrichment of complex I activity in the mitochondrial respiratory chain (GO:0032981), electron transport coupled to proton transport (GO:0015990) (Fig 4B), and the inner mitochondrial membrane (GO:0005743) (Fig 4C) further supports an adaptive mechanism aimed at enhancing energy production.

However, despite this upregulation, reductions in the efficiency of the electron transport chain (GO:0022900) (Fig 5B), ATP synthesis (GO:0042777, GO:0045259) (Fig 5C), transmembrane transport of ATPase-bound monoatomic cations (GO: 0019829), and proton-transporting ATPase activity via a rotational mechanism (GO:0046961) (Fig 5A) suggest potential mitochondrial dysfunction, possibly due to damage caused by oxidative stress [25]. This dysfunction limits the ability of cells to produce ATP efficiently, creating a detrimental cycle in which energy demand increases while production remains compromised.

Additionally, a decrease in NAD+ kinase activity (GO:0003951) (Fig 5A) was observed. This enzyme is critical for converting NAD+ to NADP+, which is an essential precursor for NADPH synthesis. NADPH serves as a crucial electron donor in biosynthetic processes and plays a vital role in antioxidant defence [53]. Although cells require both NAD+ and NADPH to maintain proper function, inflammatory stress can disrupt the balance of these coenzymes, compromising the ability of cells to fight oxidative stress, increasing inflammation, and altering cellular metabolism.

## 5. Alterations in protein synthesis and modification

During the onset of UC, a decrease in the activity of several key enzymes involved in protein biosynthesis and regulation is observed, including alanine-tRNA ligase (GO: 0004813), RNA methyltransferase (GO:0008173) (Fig 5A), alanyl-tRNA aminoacylation (GO:0006419), and lysyl-tRNA aminoacylation (GO:0006430) (Fig 5B). This decline indicates a diminished capacity to maintain protein synthesis [54]. Additionally, a decrease in serine-type endopeptidases (GO:0004252) and cysteine peptidases (GO:0008234) (Fig 5A), which are essential for the maintenance and recycling of cellular proteins, suggests an impaired ability to manage damaged proteins and maintain cellular functionality ([55], [56]).

In contrast, lysine-tRNA ligase activity (GO:0004824) (Fig 4A) is enriched at disease onset. This enzyme not only plays a crucial role in protein translation but can also act as an immune signalling molecule, activating monocytes/macrophages when secreted under certain conditions, including during inflammatory processes [57].

The mRNA polyadenylation and cleavage complex (GO:0005847) (Fig 4C) is also enriched at early stages of UC. This complex ensures proper mRNA processing, which is critical in an inflammatory context where precise regulation and stabilisation of specific mRNAs are required [58].

Furthermore, beta-galactosidase activity (GO:0004565) and beta-glucosidase activity (GO:0008422) (Fig 4A) are more enriched at UC onset compared to the control. This finding contrasts with previous studies by Mroczyńska et al. [59] and Rhodes et al. [60], who found no significant correlation between these enzymatic activities and the disease, observing similar levels among patients with UC, CD, and healthy controls. The discrepancy between these results may be attributed to factors specific to the experimental context, such as sample composition or methodology.

## 6. Alterations in transporter activity

Regarding transport mechanisms, the activity of the transmembrane phosphate transporter (GO:0005315) (Fig 5A) is less enriched in early UC than in control individuals. Acute inflammation leads to dysregulation of transporter proteins, resulting in reduced efficiency of lipid transport, an essential process for multiple cellular functions ([61], [62]). This decrease may reflect an adaptive mechanism by which intestinal cells downregulate metabolic activity to prioritize emergency energy and cellular repair processes [62].

The observed reduction in sodium ion transport (GO:0006814) (Fig 5B) may be related to the characteristic diarrhoea seen in IBM. Sullivan et al. [63] reported that diarrhoea in IBD is associated with the downregulation of various sodium transporters and regulatory proteins in the intestinal mucosa. A decline in sodium transporter activity could impair nutrient and ion absorption by cells, disrupt cellular homeostasis, and lead to excessive fluid loss in the intestine, ultimately contributing to diarrhoea, a common symptom of UC.

Additionally, the decrease in axonemal microtubules (GO:0005879) (Fig 4C), which are essential structural components for cell motility and intracellular transit, may be associated with disruptions in signal transport and intracellular trafficking in UC [64].

In contrast, the increased activity of polysaccharide transmembrane transporters (GO:0015159) (Fig 4A) at the onset of UC suggests a heightened demand for polysaccharide transport. Polysaccharides may serve as energy sources or structural components in the inflammatory response, playing a critical role in tissue repair and immune modulation, which may explain this increased activity [65].

The enrichment of xenobiotic (GO:0042908) and lipopolysaccharide (GO:0015920) transport processes (Fig 4A) indicates increased intestinal permeability and heightened exposure to toxic components. Inflammation and altered intestinal permeability facilitate the passage of external toxins and bacterial components such as lipopolysaccharides, thereby activating transport and elimination mechanisms to mitigate toxicity and manage the immune response [66]. This also reflects an environment in which dysbiosis and inflammation enhance the body’s defence response against harmful substances.

## 7. Viral involvement

The observed enrichment in viral terminase (GO:0098009) and viral envelope (GO:0019031) (Fig 4C) suggest a potential link between viral activity and the pathogenesis of UC. Emerging evidence indicates that certain viruses in the gastrointestinal tract may contribute to the development of UC by modulating the host’s immune response and promoting chronic inflammation [67]. Similarly, Li et al. [68] highlight the role of bacteriophages as natural regulators of the gut microbiota, emphasizing their ability to modulate immune responses. The complex relationship between viruses and inflammatory processes opens new avenues for research, potentially advancing our understanding of the role of viruses in UC and paving the way for novel therapeutic strategies to treat this IBD.

### 3.6 Gene Ontology: Crohn’s Disease

For CD, we followed the same approach as CU, with bar charts illustrating the up-regulated ontologies (Fig. 6) and the down-regulated ontologies (Fig. 7), both organized according to the three main GO categories.

**Fig 6.**
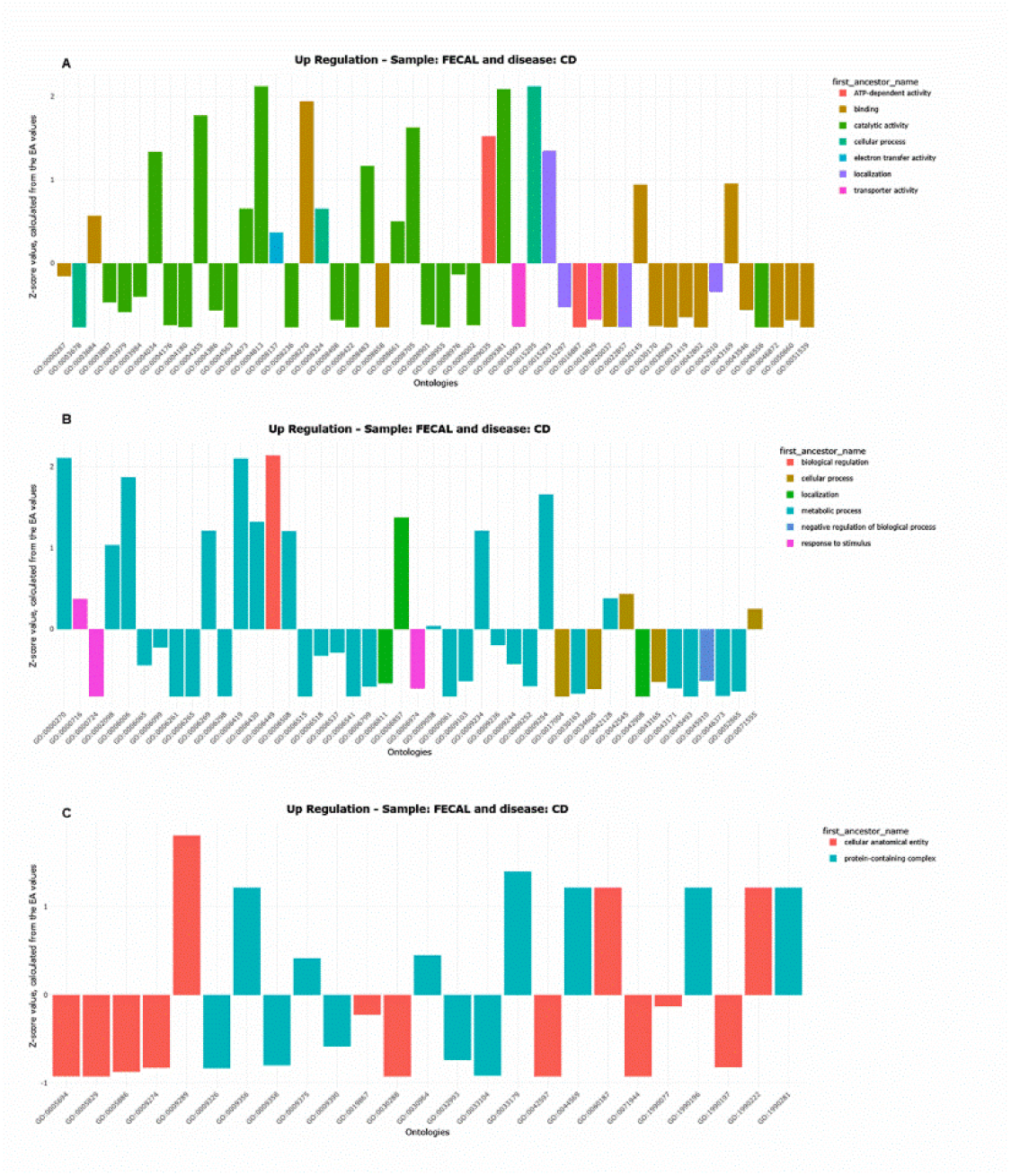
Bar Chart of Upregulated Ontologies in Crohn’s Disease. Bar chart illustrating upregulated ontologies associated with Crohn’s disease (CD). Ontologies are plotted on the X-axis, and the Z-score (derived from EA values) is plotted on the Y-axis. The Z-score normalises enrichment values, indicating how far each ontology deviates from the mean within the same disease. This normalization helps identify the most relevant ontologies by highlighting those with significantly higher or lower enrichment. Fig A represents molecular functions; Fig B, biological processes, and Fig C, cellular components, the three gene ontology categories.

**Fig 7.**
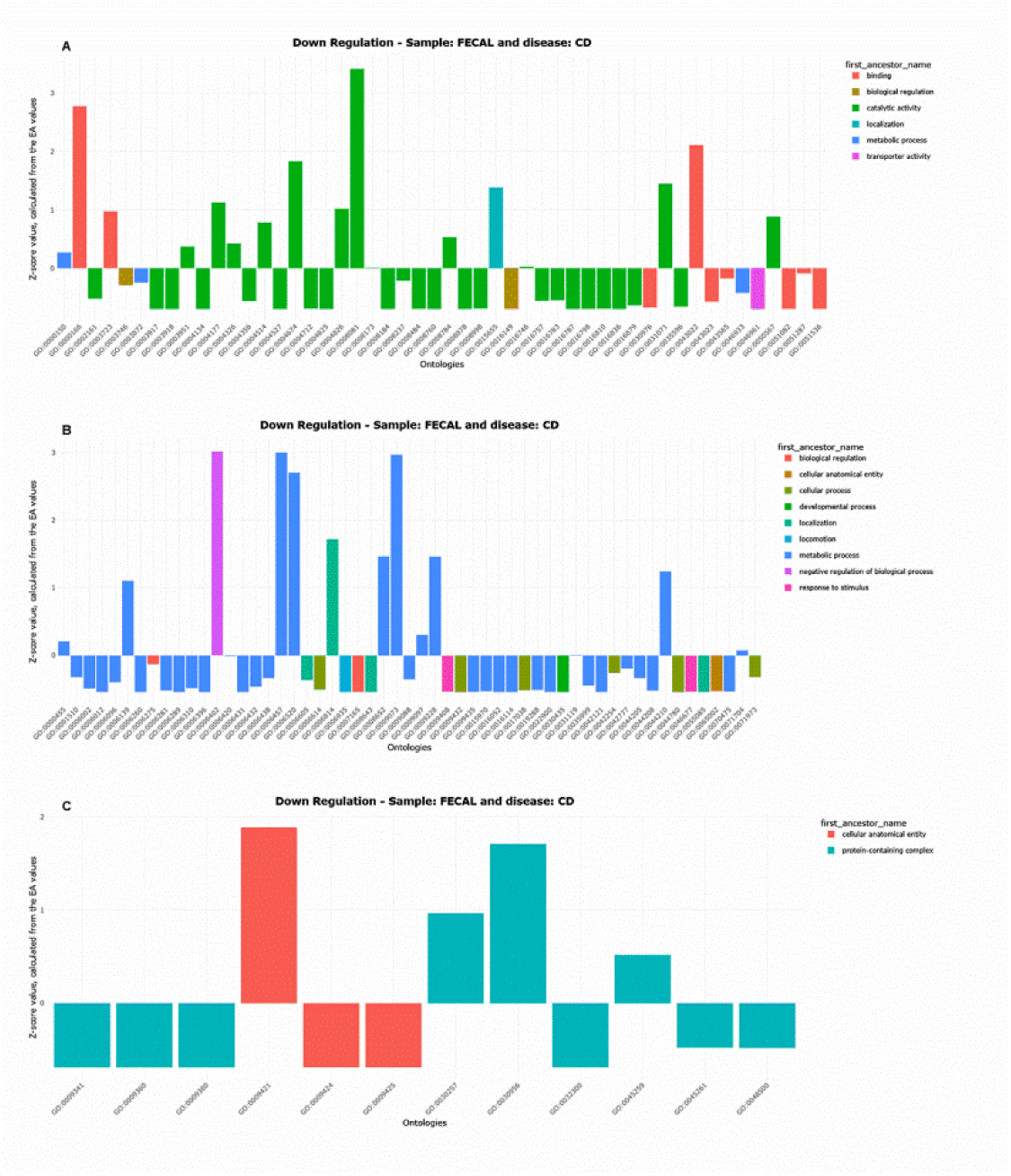
Bar Chart of Downregulated Ontologies in Crohn’s Disease. Bar chart illustrating downregulated ontologies associated with Crohn’s disease (CD). Ontologies are plotted on the X-axis, and the Z-score (derived from EA values) is plotted on the Y-axis. The Z-score normalises enrichment values, indicating how far each ontology deviates from the mean within the same disease. The normalization helps to identify the most relevant ontologies by highlighting those with significantly higher or lower enrichment. Fig A depicts molecular function; Fig B, biological processes, and Fig C, cellular components, the three main gene ontology categories.

## 1. Response to oxidative stress and cellular damage

At the onset of CD, intestinal cells face a significant increase in oxidative stress, triggering the activation of several enzymes involved in cellular repair. Notably, the enrichment of site-specific deoxyribonuclease type I (GO:0009035) (Fig 6A), which degrades damaged DNA, suggests a protective response to inflammation-induced genotoxic stress [69].

To counteract genetic damage induced by chronic inflammation and reactive oxygen species, intestinal cells may activate multiple DNA repair mechanisms [70], including homologous recombination for double-strand break repair (GO:0000724), transcription-coupled nucleotide excision repair (GO:0000716), DNA damage recognition (GO: 0000716), DNA damage response (GO:0006974) (Fig 6B), ABC excinuclease activity (GO:0009381), and DNA damage binding (GO:0003684) (Fig 6A). This coordinated response highlights the cellular effort to maintain genetic integrity amid intense oxidative stress, a key factor in the pathogenesis of CD.

Additionally, the upregulation of transmembrane nucleobase transporters (GO:0015205) (Fig 6A) facilitates the cellular uptake of adenine, guanine, cytosine, and thymine, supporting nucleotide synthesis [71]. The increase in cellular stress and activation of repair mechanisms during the early stages of CD may intensify the demand for nucleotides required for DNA and RNA synthesis, driving the activity of these transporters.

Regarding nucleotide metabolism, the reduced activity of phosphate ester hydrolases (GO:0008081) (Fig 7A), also known as phosphodiesterases (PDE) [72], may have significant implications for the regulation of inflammation and cellular function in CD. PDEs are responsible for breaking down cyclic nucleotides such as cAMP and cGMP, which are essential for cell signalling. These molecules are involved in modulating inflammation, the immune response, and other cellular processes [72].

A study by Salari-Sharif & Abdollahi [73] shows that PDE inhibitors are effective in treating inflammatory disorders. For instance, PDE4 inhibitors have been used to manage conditions such as asthma and chronic obstructive pulmonary disease, due to their ability to regulate inflammatory responses. In IBD such as CD, PDE4 inhibitors may exert anti-inflammatory, antidepressant, and antifibrinolytic effects, potentially influencing disease pathogenesis.

The reduced PDE activity observed in CD may therefore increase cAMP and cGMP levels, contributing to more efficient regulation of inflammation and mitigating excessive immune responses.

## 2. Dysbiosis and alteration of the microbiota

Chronic inflammation at the onset of CD leads to significant changes in the intestinal microbiota, characterised by shifts in bacterial populations and their associated functions. One notable alteration is the reduction in flagellar bacterial motility, as evidenced by a decrease in ontologies related to the entire bacterial flagellum (GO:0044780, GO:0071973 (Fig 7B), GO:0009424, GO:0009425, GO:0009421 (Fig 7C)). This decline may be due to the inflammatory environment, which hinders the survival and function of flagellated bacteria. As discussed in the section “Dysbiosis and Microbiota Alterations in UC”, Hsu et al. [39] demonstrated that intestinal cells in an inflamed state release proteins such as hLYPD8, which bind to bacterial flagella, limiting their mobility and preventing bacterial invasion of the epithelial barrier.

In contrast, an enrichment in pili (GO:0009289) (Fig 6C) is observed, bacterial structures that facilitate the attachment of species such as adherent-invasive *Escherichia coli* (AIEC) to intestinal cells. Pili play a crucial role in bacterial colonization and biofilm formation in the intestinal epithelium, a key factor in the development and persistence of intestinal inflammation [74].

Additionally, an enrichment of efflux pumps (GO:1990281) such as the MacAB-TolC complex (GO:1990196) and the ProVWX complex (GO:1990222) (Fig 6C) supports their importance as bacterial defence mechanisms. Efflux pumps enable bacteria to expel toxic substances, including antibiotics, heavy metals, and inflammatory metabolites, enhancing their resistance and survival in hostile environments [75].

In the context of CD, bacteria colonising the intestinal tract can activate efflux systems such as MacAB-TolC to remove harmful compounds, enhancing their resilience within the inflammatory environment [76]. The ProVWX complex, on the other hand, is enriched in CD, probably as an adaptive response to osmotic stress and chronic inflammation. In the inflamed intestinal tract, bacteria colonising the intestinal mucosa are exposed to dehydration and high solute concentrations. The upregulation of the ProVWX complex helps regulate the transport of water and solutes, facilitating the uptake of protective molecules such as betaine to mitigate osmotic damage [77]. This adaptation enables bacterial survival in the altered intestinal environment of CD, which is characterized by an overactive immune system and changes in microbiota composition.

## 3. Alterations in ribosomal activities and protein synthesis

A marked decrease in protein metabolic activities, including ribosome binding (GO:0043022), nucleotide binding (GO:0000166), RNA binding (GO:0003723) (Fig 6A), ribosome biogenesis (GO:0042254), rRNA biogenesis (GO:0000455), nucleotide synthesis (GO:0006139), and SRP-dependent cotranslational protein targeting to membrane (GO:0006614) (Fig 6B) suggests that the inflammatory environment in CD disrupts these critical cellular processes. This disruption may impair the cell’s ability to sustain protein synthesis and regeneration, both of which are essential for responding to tissue damage and inflammation.

Further supporting this trend is the decreased activity of several key enzymes involved in protein metabolism, such as aminopeptidase (GO:0004177) and phenylalanine-tRNA ligase (GO:0004826) (Fig 7A), which are less enriched at the onset of CD. Aminopeptidases play a vital role in several important processes in intestinal metabolism, including protein digestion, regulation of hormonal peptides, and the maturation of functional proteins [78]. Their reduced activity can compromise nutrient absorption, disrupt hormonal balance, and lead to the accumulation of misfolded proteins. Additionally, during the inflammatory episodes characteristic of CD, the expression of many proteins, including those involved in protein synthesis, is altered. The inflammatory response may interfere with the function of aminoacyl-tRNA synthases, such as phenylalanine-tRNA ligase, reducing their efficiency during the translation process [79].

Protein malnutrition, commonly observed in CD, is due to disruptions in essential metabolic processes related to amino acid metabolism [32]. This is evidenced by the reduced activity of amino acid metabolism (GO:0006520), amino acid biosynthesis (GO:0008652), as well as the biosynthesis of aromatic amino acids (GO: 0009073) and isoleucine (GO:0009097) (Fig 7B). These findings suggest a diminished capacity of intestinal cells to metabolize fundamental nutrients. Combined with chronic inflammation, this metabolic dysfunction, contributes to impaired amino acid absorption and decreased protein synthesis. Consequently, patients with CD are at a high risk of protein malnutrition [32], which can exacerbate clinical symptoms and negatively impact both intestinal and immune function.

In parallel, an increase was observed in ontologies related to the metabolism of specific amino acids, such as glutamate synthase activity (GO:0004355) (Fig 6A), and protein synthesis, including alanine-tRNA ligase activity (GO:0004813), transaminase activity (GO: 0008483), methionine synthase activity (GO:0008705) (Fig 6A), translation termination regulation (GO:0006449), tRNA uridine modification (GO:0002098), alanyl-tRNA aminoacylation (GO:0006419), and lysyl-tRNA aminoacylation (GO:0006430) (Fig 6B). This pattern suggests an increased demand for proteins to support immune response and tissue repair. This is particularly important in CD, where intestinal cells must sustain high levels of protein synthesis to counteract continuous mucosal damage.

It is evident that protein metabolism is altered in CD. While a general decrease in overall protein synthesis is observed, there is a selective upregulation of specific proteins, likely those involved in inflammation and tissue repair. This adaptive response may help meet the increased demands for immune response and cellular regeneration during inflammatory episodes. However, despite the clear evidence for this phenomenon, further studies are needed to elucidate the exact mechanisms underlying this selective protein synthesis and the constraints imposed by the inflammatory environment.

## 4. Dysregulation of energy pathways

Energy production is significantly impaired by inflammation at the onset of CD. This is reflected by a decrease in ATPase activity, including proton-transporting ATPase activity via a rotational mechanism (GO:0046961, GO:0046933 (Fig 7A), GO:0045259 (Fig 7C)). This decline indicates that the energetic stress induced by inflammation compromises the ability of cells to efficiently hydrolyse and synthesize ATP.

However, the observed enrichment of the V-type proton-transporting ATPase, specifically the V_0_ domain (GO:0033179) (Fig 6C), indicates that despite the overall decrease in ATP synthase activity, its subunits (V₀ and V₁) may be differentially regulated. Under inflammatory conditions, such as those present in CD, the V₀ domain, which acts as a proton channel, may be upregulated as an adaptive response to maintain proton transport. In contrast, the V₁ domain, responsible for ATP hydrolysis [80], may exhibit reduced activity or functionality. This dissociation could represent a cellular adaptation to inflammatory stress, potentially arising from post-transcriptional modifications that affect overall ATPase functionality. Although the V_0_ and V_1_ domains typically collaborate in the ATPase turnover [80], the precise details of how this process is affected under pathological conditions remain unclear. Further studies are needed to elucidate the underlying mechanisms and determine how these processes are affected in the context of IBD.

Additionally, a reduction in thiamine synthesis (GO:0009228) (Fig 7B) is a common feature in patients with digestive tract disorders, including CD. These pathologies often impair the absorption of thiamine, leading to deficiencies that affect energy metabolism [81]. As thiamine is a cofactor for key enzymes in carbohydrate metabolism and energy production, its deficiency disrupts energy production, leading to the accumulation of intermediate metabolites such as lactate. This can result in symptoms such as encephalopathy, muscle weakness, and nervous system disorders, as the nervous system is particularly sensitive to energy deprivation [81].

## 5. Regulation of cellular transport and homeostasis

The increase in transport-related ontologies, such as transport of monoatomic cations (GO:0008324) (Fig 6A), suggests a cellular effort to maintain osmotic homeostasis and function in a hostile environment [82]. Notably, the enrichment of ferrous iron transport (GO:0015093) (Fig 6A) is particularly significant. According to Marques et al. [83], during chronic inflammation, immune cells deplete oxygen in the local epithelial microenvironment, resulting in localized hypoxia. In response, regulatory mechanisms such as hypoxia-inducing factors (HIFs) enhance iron absorption by upregulating genes responsible for duodenal iron transport, including divalent metal transporter 1 (DMT1). Interestingly, Marques et al. [83] also report that certain intestinal bacteria, such as *Lactobacilli*, produce metabolites capable of modulating HIF activity, potentially inhibiting it under specific conditions. This bacterial strategy may serve to reduce intestinal iron absorption, limiting its availability to the host. Thus, the observed increase in ferrous iron transport may represent an adaptive response driven by HIFs, which upregulate genes such as DMT1 to meet iron demands while counteracting bacterial-induced iron accumulation.

Additionally, the increased transport of xenobiotics (GO:0042908) and oligopeptides (GO:0006857) (Fig 6B) suggests a heightened intestinal permeability, leading to greater exposure to toxic substances. The enrichment of oligopeptide transporters in CD may be related to the presence of proinflammatory peptides secreted by intestinal bacteria such as *Escherichia coli* [84]. These peptides can induce intestinal inflammation, as observed when Pept-1, an oligopeptide transporter, mediates their uptake and triggers an inflammatory response [84]. In IBD patients, increased expression of Pept-1 in the colon has been reported, possibly in response to elevated levels of proinflammatory bacterial peptides. This suggests that the oligopeptide transport system is activated under inflammatory conditions, with bacterial peptides serving as substrates that induce Pept-1 expression, further amplifying intestinal inflammation [84]. Finally, the upregulation of xenobiotic transporters may help eliminate toxic by-products generated during the immune response, particularly in an environment with altered microbiota, where harmful metabolites are more likely to accumulate [85].

The increase in histidine kinase activity (GO:0004673) (Fig 6A) is associated with cell signalling processes, as described by Wolanin et al. [86]. In CD, dysregulation of cell signalling may contribute to tissue damage and an exaggerated immune response, which explains why these activities are more prominent in the early phase of the disease.

Finally, according to Sullivan et al. [63], diarrhoea in IBD may be linked to the downregulation of several sodium transporters, including sodium symporters (GO:0015655) (Fig 7A) and sodium ion transporters (GO:0006814) (Fig 7B). A decrease in sodium transporter activity impairs the ability of intestinal cells to absorb nutrients and ions effectively, leading to excessive fluid loss in the intestine. This mechanism contributes to the chronic diarrhoea commonly observed in IBD patients.

### 3.7. Differences between CD and UC in Gene Ontology

Gene ontology (GO) analysis provides insights into the molecular and cellular mechanisms underlying Ulcerative Colitis (UC) and Crohn’s Disease (CD). The following table summarizes key differences in GO terms between these two inflammatory bowel diseases, highlighting variations in oxidative stress response, microbiota interactions, metabolism, and cellular transport.

## 4. Conclusions

Functional analysis in paediatric patients with CD and UC has identified significant differences in metabolic pathways and genetic ontologies, particularly related to inflammation, dysbiosis, and immune function. These findings suggest that each condition involves distinct molecular mechanisms and highlight the need for further research into the role of the intestinal microbiota in IBD. Notably, contradictory / divergent results in KEGG pathway expression and protein synthesis alterations revealed by GO term analysis remain unexplained. Although this study is exploratory in nature and based on a small sample of 32 patients, its findings may be relevant for the development of new diagnostic and therapeutic strategies. However, larger-scale studies are necessary to validate and generalize the results.

## 5. Data availability statement

The datasets presented in this study can be found in online repositories. The names of the repository/repositories and accession number(s) can be found in the article/Supplementary Material.

## Data Availability

The raw data from which this analysis was carried out can be found in a repository

https://doi.org/10.34810/data593

## Data Availability

The raw data from which this analysis was carried out can be found in a repository

https://doi.org/10.34810/data593

## 6 Acknowledgments

Our thanks to Mrs Lucy Brzoska for reviewing the text.

## 8. Supplementary Material

The Supplementary Material for this article can be found online at: https://www.frontiersin.org/articles/10.3389/fped.2023.1220976/full#supplementary-material

## 9. Ethics statement

The studies involving humans were approved by the Ethical Committee of the Sant Joan de Déu Hospital (Esplugues de Llobregat, Barcelona). The studies were conducted in accordance with the local legislation and institutional requirements. Written informed consent for participation in this study was provided by the participants’ legal guardians/next of kin.

This specific study represents a continuation and focused extension of a larger, published [11] pre-existing study. Its primary objective is to complete and interpret only a subset of the extensive data and results previously obtained. Crucially, no additional biological or bioinformatic analyses were performed for the current manuscript; the work is focused solely on the interpretation of previously secured data and outcomes.

